# A multimodal neuroimaging study of brain abnormalities and clinical correlates in post treatment Lyme disease

**DOI:** 10.1101/2022.07.01.22277150

**Authors:** Cherie L. Marvel, Kylie H. Alm, Deeya Bhattacharya, Alison W. Rebman, Arnold Bakker, Owen P. Morgan, Jason A. Creighton, Erica A. Kozero, Arun Venkatesan, Prianca A. Nadkarni, John N. Aucott

## Abstract

Lyme disease is the most common vector-borne infectious disease in the United States. Post-treatment Lyme disease (PTLD) is a condition affecting 10-20% of patients in which symptoms persist despite antibiotic treatment. Cognitive complaints are common among those with PTLD, suggesting that brain changes are associated with the course of the illness. However, there has been a paucity of evidence to explain the cognitive difficulties expressed by patients with PTLD.

This study administered a working memory task to a carefully screened group of 12 patients with well-characterized PTLD and 18 healthy controls while undergoing functional MRI (fMRI). A subset of 12 controls and all 12 PTLD participants also received diffusion tensor imaging (DTI) to measure white matter integrity. Clinical variables were also assessed and correlated with these multimodal MRI findings.

On the working memory task, the patients with PTLD responded more slowly, but no less accurately, than did controls. FMRI activations were observed in expected regions by the controls, and to a lesser extent, by the PTLD participants. The PTLD group also hypoactivated several regions relevant to the task. Conversely, novel regions were activated by the PTLD group that were not observed in controls, suggesting a compensatory mechanism. Notably, three activations were located in white matter of the frontal lobe. DTI measures applied to these three regions of interest revealed that higher axial diffusivity correlated with fewer cognitive and neurological symptoms. Whole-brain DTI analyses revealed several frontal lobe regions in which higher axial diffusivity in the patients with PTLD correlated with longer duration of illness. Together, these results show that the brain is altered by PTLD, involving changes to white matter within the frontal lobe. Higher axial diffusivity may reflect white matter repair and healing over time, rather than pathology, and cognition appears to be dynamically affected throughout this repair process.

## Introduction

Lyme disease is a vector-borne infectious disease initiated by the bite of a tick infected with various genospecies of the bacteria *Borrelia burgdorferi* sensu lato [1]. In recent decades, both the geographic range and the number of cases have increased significantly, and the Centers for Disease Control and Prevention (CDC) recently estimated an incidence of 476,000 cases annually in the US [2]. Untreated Lyme disease can manifest clinically as the skin rash erythema migrans, or cause cardiac, neurologic, or joint signs of infection resulting from the dissemination of the bacteria [1, 3]. Lyme disease is treated with antibiotics, after which such symptoms typically resolve.

However, a subset of patients (10-20%) who are appropriately treated for Lyme disease develop a chronic illness consisting of persistent or recurrent symptoms [4, 5]. A specific, research-based definition for post-treatment Lyme disease (PTLD) has been operationalized to identify patients with symptoms linked temporally to strong evidence of prior exposure to *B. burgdorferi* [5–7]. Although fatigue, widespread musculoskeletal pain, and cognitive difficulties are the most prominent symptoms of PTLD, patients also often report a constellation of other neurologic, sleep, ocular, mood, and other symptoms [7–9]. Symptom severity and course can be variable, yet PTLD often significantly impacts cognition and health-related quality of life [7, 10–13]. There is currently no sensitive or specific test to aid diagnosis of PTLD, nor are there FDA-approved treatment options for patients. Research in individuals suffering from PTLD has been relatively sparse, in part, due to the complexity of the disease and the difficulty in confirming a PTLD diagnosis in the absence of additional underlying or co-morbid diseases that would complicate the interpretation of research results.

The underlying mechanisms of brain changes that may impact cognition in people with PTLD are largely unknown. Few neuroimaging studies that have been reported in people with PTLD, and there is a lack of consistent findings explaining neurological deficits [14–17]. Several brain perfusion and metabolism studies have shown abnormal patterns in patients who underwent antibiotic treatment [14–17]. It has been noted that brain changes associated with Lyme disease may involve abnormal white matter perfusion that impacts cognition [18, 19]. Moreover, microglial activation of patients with PTLD has been suggested as a contributing factor of PTLD-related neurological deficits [20]. However, state-of-the-art magnetic resonance imaging (MRI) methods that measure the structural and functional integrity of gray and white matter in PTLD have not been reported to date, limiting our understanding of neurologic deficits related to PTLD.

The aim of the current study was to use functional magnetic resonance imaging (fMRI) and diffusion tensor imaging (DTI) methods to examine brain function and structure in people with PTLD. We sought to test the hypothesis that people with PTLD show altered task-related activations as revealed by fMRI, and white matter abnormalities as revealed by DTI.

## Materials and methods

### Participants

Twelve adult participants with PTLD (≥ 18 years) were originally recruited from a referral-based clinic population. Those providing permission to contact for future studies as part of the consent process were approached for the current MRI study. The median time between participation in both studies was approximately 2.5 weeks (range <1 week – 9.4 weeks).

Study participant selection methods replicated many of the criteria set forth in the Infectious Diseases Society of America’s (IDSA) proposed case definition for PTLD [6]. A rigorous chart review process confirmed study eligibility to validate PTLD in the absence of confounding factors. Confirmation of PTLD was determined by: 1) physician-documented erythema migrans rash, or 2) evidence of new-onset objective signs (e.g., joint swelling, facial palsy) <u>and</u> laboratory evidence of infection following CDC recommendations for test interpretation, or 3) evidence of new-onset symptoms not attributable to another cause <u>and</u> laboratory evidence of infection following CDC recommendation for test interpretation. Along with PTLD confirmation, eligibility required a history of appropriate antibiotic treatment and post treatment symptoms specified in the IDSA case definition; fatigue, musculoskeletal pain, and/or cognitive difficulty. Additionally, at least one symptom had been experienced in the past two weeks that limited daily functioning at least half the time when present. Further eligibility inclusion and exclusion criteria for this study have been published previously [7].

Participants were excluded for a range of specific, co-morbid conditions with significant symptom overlap with PTLD such as fibromyalgia, chronic fatigue syndrome, major psychiatric disease conditions (except non-suicidal depression that manifested after Lyme infection), malignancy, and autoimmune disease. Exclusion criteria also consisted of: history of Lyme vaccine, sleep apnea, cirrhosis, hepatitis B/C, HIV, dementia, cancer (past 2 years), illicit substance abuse, prescription drug abuse, and alcoholism.

A total of 18 adult control participants were recruited through community flyers and included in the fMRI analysis. Control participants were additionally screened for any co-morbid conditions with significant symptom overlap with PTLD, exclusion criteria as described above, or a past diagnosis of Lyme disease.

In a final screening stage, both participants with PTLD and controls were excluded from study participation if they endorsed the following characteristics that might confound data interpretation: major neurologic disorders (including stroke and seizures); head injury resulting in loss of consciousness of > 5 minutes, significant learning disability, left-handedness, or being a non-native English speaker (i.e., acquired English post puberty). Participants were also excluded for reasons of safety concerns within the MRI environment, such as: current or possible pregnancy, metal inside or attached to the body, and claustrophobia.

Demographic and clinical characteristics are summarized in Table 1. A subset of 12 controls who were matched demographically to the PTLD group also had DTI data available and were included in the DTI analysis.

**Table 1.** Baseline demographic and clinical characteristics. N (%) are presented for categorical variable; mean (standard deviation), 95% confidence interval [lower limit, upper limit] are presented for continuous variables; Shapiro-Wilk tests were used for tests of normality. Significant values of the Shapiro-Wilk tests are denoted in bold, p ≤.05, two-tailed. Control groups did not differ from PTLD for age, education, or gender. ^a^Total days from Lyme disease onset (or start of antibiotics) until MRI scan ^b^The total number of symptoms reported at the moderate or severe level over the past two weeks on the Post-Lyme Questionnaire of Symptoms (range of 0 to 36). ^c^Two-tier tests were interpreted using CDC criteria for positivity, which incorporates duration of illness at the time of the test. ^d^Confirmed through medical record review. Participants presenting with erythema migrans rash were not required to have a concurrent positive two-tier serology. Those with neurologic disease (*n*=2 with Bell’s Palsy, *n*=1 with meningitis/encephalitis), late Lyme arthritis, or an initial flulike illness were required to have a concurrent positive two-tier test. ^e^Beck Depression Inventory: Cognitive/Affective Subscale score

| Group | PTLD<br>*n* = 12 | Controls<br>(for fMRI)<br>*n* = 18 | Matched Controls<br>(for DTI)<br>*n* = 12 |
| --- | --- | --- | --- |
| Age (years) | 45.16 (13.62)<br>[36.51, 53.82], *W* = .96, *p* =<br>.72 | 47.01 (13.10)<br>[40.49, 53.52], *W* = .86, *p* =<br>.01 | 45.33 (13.76)<br>[36.60, 54.08], *W* = .91, *p* = .20 |
| Male Gender | 7 (58.33%) | 6 (33.33%) | 4 (33.33%) |
| Education (years) | 16.17 (2.21) | 16.44 (2.01) | 16.00 (1.91)<br>[14.79, 17.21], *W* = .86, *p* = .05 |
|  | [14.76, 17.57], W = .97, p =<br>.92 | [15.45, 17.44], W = .90, p =<br>.06 |  |
| Duration of illness<sup>a</sup> (days) | 944.83 (1043.13)<br>[282.06, 1607.61], W = .75, p<br>= **.003** |  |  |
| Antibiotic exposure<sup>a</sup><br>(days) | 190.17 (309.37)<br>[-6.40, 386.73], W = .55, p <<br>**.001** |  |  |
| Number of symptoms at<br>MRI scan<sup>b</sup> | 8.17 (4.39)<br>[5.38, 10.95], W = .94, p = .48 |  |  |
| Neurologic symptoms | 4.50 (2.02<br>[3.21, 5.79], W = .90, p = .16 |  |  |
| Cognitive symptoms | 2.00 (1.35)<br>[1.14, 2.86], W = .71, p = **.001** |  |  |
| Two-tier antibody<br>positive at MRI scan<sup>c</sup> | 4 (33.33%) |  |  |
| Initial Lyme disease clinical presentation<sup>d</sup> |  |  |  |
| Erythema migrans rash | 4 (33.33%) |  |  |
| Neurologic Lyme<br>disease | 3 (25.00%) |  |  |
| Late Lyme arthritis | 1 (8.33%) |  |  |
| Flulike illness | 4 (33.33%) |  |  |
| Beck Depression Inventory<sup>e</sup> | 18.42 (12.05)<br>[10.76, 26.07], W = .79, **p = .008** |  |  |

The Institutional Review Board of the Johns Hopkins University School of Medicine approved this study, and written informed consent was obtained from all study participants prior to initiation of study activities. The planning and conduct of this research were in accordance with the Helsinki Declaration as revised in 2013.

### Clinical data collection

Participants were asked to complete a self-administered 36-item post-Lyme questionnaire of symptoms (PLQS) which was developed based on prior clinical and research experience among patients with PTLD. The list of individual symptom items has been previously published [22]. For each item, participants indicated severity over the past 2 weeks (0=absent, 1=mild, 2=moderate, 3=severe), and a binary response was created (absent/mild =0 vs. moderate/severe = 1). A total symptom score was generated by summing each binary item (range 0-36). Additionally, in order to focus on specific symptoms of interest to the current study, a ‘neurologic’ symptom score was generated a priori by summing the binary responses for the following 12 symptoms; fatigue, numbness in hands/feet, numbness in face/scalp, headache, photophobia, drooping facial muscle, drooping eyelid, neck pain, poor coordination, memory impairment, difficulty finding words, and difficulty focusing or concentrating (range 0-12). Finally, a more narrow ‘cognitive’ symptom score was generated by summing the binary responses for the following 3 items only; memory impairment, difficulty finding words, and difficulty focusing or concentrating (range 0-3).

### MRI procedures

#### Behavioral Task

Participants were asked to perform a working memory task in the MRI scanner consisting of two conditions, previously described in detail [23, 24]. Briefly, in the control condition, participants viewed one or two uppercase consonants (one second), followed by a blank screen (four or six seconds).

Participants held these letters in mind through silent rehearsal. Finally, a single lowercase letter was presented (one second). Participants indicated via button press whether the single probe item matched either of the targets presented at the start of the trial. In the “forward” condition, rather than rehearse the original target letters presented at the start of the trial, participants were required to count two alphabetical letters forward of each target letter(s) and hold the new letters in mind. For example, if the target letters were ‘‘f’’ and ‘‘q’’, participants would count forward to the letters ‘‘h’’ and ‘‘s’’. When the probe letter appeared, participants indicated whether the probe matched the newly derived letters instead of the original target letters. Therefore, the two conditions differed specifically during the rehearsal phase of the trial, in which target letters were simply rehearsed as presented (control condition) or rehearsed by counting two alphabetical letters forward (forward condition).

Participants were instructed to respond as quickly and accurately as possible while completing all operations silently ‘‘in your head.’’ Button press responses were recorded if conducted within six seconds following probe onset (yes = right index finger; no = right middle finger). Trials were jittered with an inter-trial interval (ITI) of six to nine seconds. Response time (RT) and accuracy were recorded for each trial. In order to familiarize subjects with the rules of the task, subjects practiced 10 trials of each condition prior to entering the MRI environment.

The control and forward conditions were completed during two separate blocks, with the order counter-balanced across participants. Each block contained 64 trials (∼ 16 minutes). Each trial consisted of pseudorandom presentations of targets such that letters were unique within a trial. Probes matched a target (or newly derived target) on 50% of the trials. The number of target letters (1 or 2), rehearsal duration (4 or 6 seconds), expected response (yes or no), and duration of ITI (6 – 9 seconds) were pseudorandomized so that presentation of identical parameters was limited to three consecutive trials.

An additional event-related finger tapping task was administered in order to compute individualized hemodynamic response functions (HRFs) [23, 25]. This task consisted of a button press with the right index finger every 29-31 seconds upon presentation of a 1-second cue to “tap” followed by “rest”, which lasted for 10 minutes total. The individualized HRFs were used in the convolution step of MRI processing, rather than a canonical HRF, in case the HRFs in the PTLD group differed from that of control participants.

Stimuli were delivered using E-Prime 2.0 software (Psychology Software Tools, Pittsburgh, PA) on a Dell Optiplex SX9202 workstation running Windows 7. The stimuli were rear-projected onto a screen in the MRI scanner, which was then reflected into a head coil-mounted mirror within the participant’s line of sight. Responses were collected using two fiber optic button boxes (MRA, Inc., Washington, PA) that were held in the participant’s right hand.

#### MRI data acquisition

MRI data were acquired on a Philips 3 Tesla scanner using a 32-channel head coil. Structural images were collected using a sagittal magnetization prepared gradient-echo (MPRAGE) sequence aligned to the anterior-posterior commissure (AC-PC) axis: repetition time (TR)/echo time (TE) = 6.9/3.3 ms; field of view = 240 x 240; 170 slices; slice thickness 1.0 mm; 0 mm gap; flip angle = 8 degrees; voxel size = 0.75 x 0.75 x 1.0 mm. The total scan duration was 6 minutes. FMRI data were collected using a T2-weighted gradient echo EPI pulse sequence (TR = 1000 msec; TE = 30 ms; flip = 61; in-plane resolution = 3.75 mm; slice thickness = 6 mm with a 1 mm gap; 20 oblique-axial slices; FOV = 240 mm). T2-weighted images were acquired in the oblique-axial plane rotated 25 degrees clockwise with respect to the AC-PC line in order to optimize imaging of the cerebellum and neocortex. The number of acquired volumes within each block ranged from 917 to 922 for the working memory tasks and 600 for the tapping task. The start of the fMRI scan was triggered by E-prime software at the beginning of each block.

#### Functional MRI data analysis

The SPM12 software package (Wellcome Department of Cognitive Neurology) was used for preprocessing and statistical computations. High temporal resolution fMRI in conjunction with neocortical-specific HRFs were used to ensure maximum accuracy in characterizing phase-specific blood oxygen level dependent (BOLD) responses [23, 25]. Individual HRF regressors were convolved with reference waveforms for the target encoding (1 sec), rehearsal (4 or 6 sec) and probe retrieval (6-9 sec) phases of the task for each subject within the first-level event-related analysis. In this report, we only focus on the rehearsal phase of analyses. Standard image preprocessing steps were performed, including slice timing correction (reference = middle slice), motion correction, anatomical co-registration, normalization to the Montreal Neurological Institute (MNI) stereotaxic space, and spatial smoothing (FWHM = 8 mm). Due to a technical error, one PTLD participant’s functional MRI data was corrupted and could not be processed. It was excluded from the functional imaging stages of analysis. Individual statistical maps were computed for each subject using the general linear model approach as implemented in SPM12, with high pass filtering of 128 s. A random effects analysis was then performed to map the average responses to the rehearsal phase of the task on correct trials only. Incorrect trials were not given a regressor and were considered as residual variance. This analysis was performed by computing a contrast volume per subject and using these volumes to calculate one-sample t-test values at every voxel. Of particular interest were within-group contrasts comparing the BOLD signal difference between the 2-target forward minus 2-target control working memory conditions that were then compared between groups. MNI coordinates were transformed into the coordinate system of the Talairach and Tourneaux stereotaxic atlas [26] using the MNI to Talairach transformation described by Lancaster *et al*. [27] in order to make anatomical determinations of the activations. However, MNI coordinates are reported in the tables and figures. Significance levels were set to *p* < .005, uncorrected, with a minimum cluster size of 10 voxels.

Functionally defined regions of interest (ROIs) were circumscribed on each participant’s scan based on the activation clusters observed in the between-groups contrast using the MarsBaR toolbox for SPM [28]. The resultant contrast values per participant were then entered into subsequent analyses to test for correlations with behavioral task performance and diffusion weighted imaging (DTI) measures.

#### DWI data acquisition and preprocessing

Abnormal white matter fMRI findings led us to investigate the relationship to white matter structural integrity by adding diffusion tensor imaging (DTI) methods to the protocol already in progress, which was administered to a subset of participants (*n* = 12 per group).

Diffusion-weighted images (DWI) were acquired using a spin echo sequence with TR = 7012 ms, TE = 75 ms, FOV = 212 x 212 mm^2^, 0.83 x 0.83 x 2.2 mm voxels, flip angle = 90°, b-value = 700, number of gradients = 33, and 70 axial slices. Two sequences were acquired. The DWI data were preprocessed using FSL [29] to correct for eddy current-induced distortions and subject motion using affine registration. The b-vector matrix was adjusted based on rigid body registration and skull stripping was performed using FSL’s automated brain extraction tool (BET) to remove non-brain tissue. A standard least squares diffusion tensor fitting model was applied to the data to derive whole brain maps for the following diffusion tensor imaging (DTI) metrics: fractional anisotropy (FA), mean diffusivity (MD), radial diffusivity (RD), and axial diffusivity (AD). These estimates were computed on a voxel-by-voxel basis using a three-dimensional Gaussian distribution model that yielded a single mean ellipsoid for each voxel. For each participant, the two runs of DWI data were preprocessed separately, and the scalar maps resulting from each run were averaged to improve signal-to-noise ratio.

#### Tissue class segmentation analysis

To determine the proportion of white matter within each significant cluster of activation derived from the fMRI analysis, a tissue class segmentation analysis was used. First, a study-specific T1 modal model template was created from all participants in the study using Advanced Normalization Tools (ANTs), and a 3D vector field transformation for each subject was calculated to align the individual’s structural scan to the template modal model based on the entire sample [30, 31]. Tissue class segmentation analysis was then completed on the template modal model using FSL’s FAST automated segmentation tool [32], a well-validated automated tissue segmentation tool [33]. FAST utilizes a hidden Markov random field model and an associated expectation-maximization algorithm to segment brain images into three tissue classes: gray matter, white matter, and cerebrospinal fluid. To compute the overlap between the resulting binary white matter segmentation mask and each significant functional activation cluster, each cluster of activation was first transformed into a binary ROI. The binary ROIs were subsequently resampled into a 1 x 1 x 1 mm space using nearest neighbor interpolation, given that both the tissue class segmentation analysis and the DTI analyses were completed in this space. Following registration, each functional ROI mask was multiplied by the binary white matter segmentation mask to identify voxels in each ROI that fell within the white matter segmentation mask. The number of voxels in the subsequent overlap image was then calculated as a percentage relative to the ROI size, yielding the percentage of voxels within a particular ROI that were classified as white matter. This method was repeated for each of the significant activation clusters from the fMRI analysis. ROIs classified as greater than 50% white matter were selected for further analysis.

#### DTI data analysis

FSL’s Tract-based Spatial Statistics (TBSS) pipeline [34] was used to derive a mean white matter skeleton from participants’ DTI data that represents the center of all white matter tracts common to the sample. First, each participant’s FA map was registered to every other participant’s FA map using the nonlinear registration tool, FNIRT [35, 36]. The “most representative” image, i.e. the image that required the least warping to align every other image to it, was chosen as the target for registration and affine-transformed into MNI152 standard 1 x 1 x 1 mm space. Then, all other FA maps were transformed into this standard space by combining the individual nonlinear transforms to the target FA map with the affine transform from the target to MNI space. Next, the co-registered FA maps were merged into one 4D image, where each volume is a specific participant’s standard space FA map. From this 4D image, the mean FA image was computed and thinned using an FA threshold of 0.2, which retained only the center of all fiber pathways common to the group, generating a mean white matter skeleton. This skeletonization process ensures that subsequent analyses are restricted to tracts that are well-aligned across participants, thereby reducing potential misregistrations as a source of false positives. It also ensures subsequent analyses are less susceptible to partial volume effects. Finally, each participant’s aligned FA map was projected onto the mean skeleton to generate skeletonized FA data for each participant. The previously computed warps and skeleton projections were also applied to MD, RD, and AD maps in order to align them into MNI152 1 x 1 x 1 mm standard space and create participant level skeletonized MD, RD, and AD data.

The entire TBSS process was repeated for a patient-only whole brain analysis to examine whether voxel-wise DTI metrics correlated with duration of illness (DOI) in the patients with PTLD. We held an agnostic interest in DOI due to the practical question of whether longer exposure to PTLD led to relevant brain changes. The TBSS processing steps were the same as detailed above, with only the PTLD patient group included in this iteration. The resulting skeletonized DTI data were used to generate voxel-wise cross-subject statistics using “randomise” [37], FSL’s tool for nonparametric permutation inference testing (using 2000 permutations). GLM contrasts were constructed to test for both positive and negative correlations between the voxel-wise skeletonized DTI data and the DOI variable. Family-wise error (FWE) correction was performed using threshold-free cluster enhancement [38], which avoids the use of an arbitrary threshold for the initial cluster formation. A *p*-value < .05, FWE-corrected for multiple comparisons was considered statistically significant. For visualization of results, the “tbss_fill” script was used to enhance ease of viewing.

Mean white matter microstructure was computed for the functional activation ROIs that consisted of greater than 50% white matter, as determined by the tissue class segmentation analysis. Using each binarized functional activation ROI as a mask, mean FA, MD, RD, and AD values were extracted from each individual participant’s skeletonized DTI data.

To compute the percentage of overlap between the functional activation ROIs and the white matter skeleton, each binarized functional activation ROI mask was multiplied by the mean white matter skeleton generated from the TBSS pipeline to isolate the voxels from the functional ROIs that fell within the white matter skeleton. The number of overlapping voxels for each ROI was then computed, and results were expressed as a percentage relative to the ROI size. This analysis was only computed for ROIs consisting of greater than 50% white matter, as determined by the tissue class segmentation analysis.

To investigate the white matter surrounding the frontal lobe activation ROIs, we sought to quantify overlap between the observed functional ROIs and long-range white matter pathways. For this analysis, we examined the 42 standard white matter tracts generated from the XTRACT atlas [39]. Each tract mask was binarized and then multiplied by each binarized functional activation ROI to isolate ROI voxels that fell within the associated long-range white matter tract. The percent overlap for each ROI with each long-range tract was then computed and expressed as a percentage relative to ROI size.

#### Statistical analyses of clinical and behavioral variables

The clinical, behavioral, MRI, and DTI data collected in this study contained continuous variables, with the exception of gender (categorical data). T-tests were used to compare continuous variables (e.g., age and education), and Pearson chi-square tests were used to compare categorical data (e.g., gender). Shapiro-Wilk tests were used to determine if continuous variables followed a normal distribution. If a variable was not normally distributed, Mann-Whitney U tests were conducted to compare groups. Mixed-design ANOVAs were used to compare repeated measures between the two groups (e.g., fMRI task performance). [No ANOVAs contained a within-subjects factor with more than two levels; sphericity corrections were, therefore, not needed.] Pearson correlations were used when the Shapiro-Wilk’s normality tests indicated a normal data distribution (e.g., correlating fMRI data with the PLQS). Otherwise, Spearman’s rho non-parametric correlations were used. All tests were two-tailed, with an alpha level ≤ .05 to define statistical significance. Statistics were performed using IBM SPSS Statistics, Macintosh, version 27.0 (IBM Corp., Armonk, NY, USA).

### Data availability

All data and analysis code are available on the Open Science Framework (https://osf.io/kshq7).

## Results

The total healthy control group did not differ from the PTLD group in terms of age, (*Mdn* = 51.1), *U* = 101, p = .76, *r* = 18.4, or education level, t(22) = .35, *p* = .730, *d* = .133. The matched control group also did not differ from the PTLD group in terms of age, t(22) = .03, *p* = .975, *d* = .013, or education level, (*Mdn* = 16.0), *U* = 74.0, p = .91., *r* = 15.1. Pearson’s Chi Square tests were used to determine that gender counts also did not differ between the healthy control and PTLD groups, χ^2^(1, 30) = 1.83, *p* = .176, or between the matched control and PTLD groups, χ^2^(1, 24) = 1.51, *p* = .219.

### Behavioral results

Mean accuracy and RT (for accurate trials only) were computed for the following trial types: control condition, 1 stimulus; control condition, 2 stimuli; forward condition, 1 stimulus; and forward condition, 2 stimuli for each group. Due to a technical error, we were unable to collect behavioral data from one PTLD participant, which also removed them from the fMRI analyses, but their data were included in the DTI analyses. (S3 File Table 1) A 2(condition: control vs. forward) x 2(stimulus number: 1 vs. 2) x 2(group: controls vs. PTLD) mixed-design ANOVA yielded a main effect of condition *F*(1, 27) = 6.51, *p* = .017, η_p_^2^ = .019, and stimulus number *F*(1, 27) = 20.6, *p* < .001, η_p_^2^ = .43, indicating that participants’ accuracy decreased as a function of higher working memory load requirements. This was confirmed by an interaction of stimulus number x condition, *F*(1, 27) = 13.9, *p* = .001, η_p_^2^ = .34, indicating that trials were least accurate in the forward, 2 stimuli trial type. There were no main effect or interactions involving group, all *p*-values > .416 (Fig 1A). A 2(condition) x 2(stimulus number) x 2(group) mixed-design ANOVA was also conducted for the RT measure. As with the accuracy measure, there were main effects of condition, *F*(1, 27) = 51.3, *p* < .001, η_p_^2^ = .66, and stimulus number, *F*(1, 27) = 60.9, *p* < .001, η_p_^2^ = .69. There was also an interaction of condition x stimulus number, *F*(1, 27) = 35.3, *p* < .001, η_p_^2^ = .57. There were no interactions involving group. However, there was a main effect of group, showing that the PTLD group responded more slowly overall than did controls, *F*(1, 27) = 4.80, *p* = .037, η_p_^2^ = .15 (Fig 1B). Thus, participants found the forward, 2 stimuli trial type to be disproportionately more difficult than other trial types, as evidenced by decreased accuracy and slowed RTs. Moreover, the PTLD group showed general motor slowing.

**Fig 1.**
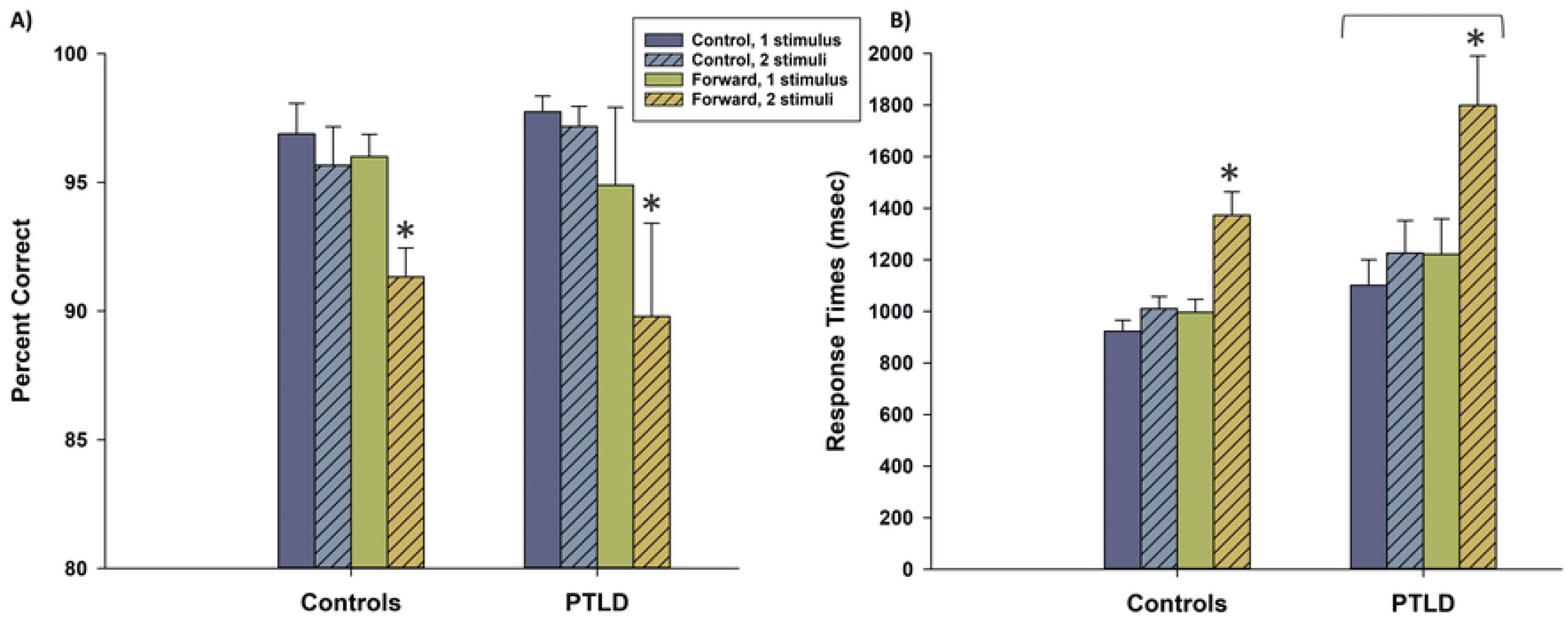
Behavioral performance on the fMRI working memory task. (A) Accuracy performance is presented, broken down by condition (control vs. forward), stimulus number (1 vs. 2 letters), and study group. Accuracy was particularly low for participants in the forward condition with two letters. However, accuracy performance did not differ overall between groups. (B) Response times for accurate trials are presented, as in (A). Response times were slowest in the forward condition with two letters. Overall, the PTLD group responded more slowly than did controls, as indicated by the overhead bracket. * indicates condition x stimulus interaction, *p* < .001; bracket indicates overall group difference, *p* = .037. Error bars denote one standard error.

### Functional MRI results

Functional imaging analyses focused on the rehearsal phase of each trial. Within this phase, we focused on activations during the most difficult condition (2 stimuli, forward condition). To do so, we computed the contrast values of the “2-target forward” minus “2-target control” conditions for each participant. To validate the results of our task, we first examined the healthy control fMRI data and compared it to a prior study that characterized this task in young, healthy adults [23] (S1 Table). Results generally overlapped with those original findings (the current study included healthy participants who were about 20 years older), with increased BOLD signal in association with verbal working memory rehearsal in the frontal lobe (BA 9 and 32), premotor cortex, caudate, thalamus, inferior parietal lobe, and superior cerebellum. The large degree of overlap with the original study supported the fMRI task’s validity. The PTLD group also revealed regions of overlap with those original findings (S2 Table), with increased BOLD signal in the frontal lobe (BA 9, supplementary motor area, and left inferior frontal gyrus BA 45), premotor cortex, caudate, and precuneus. A number of activated regions, however, were observed in the PTLD group but not observed in the controls’ data or in the original study. This suggested that the PTLD group was unable to fully utilize a typical verbal working memory circuit and compensated to maintain high accuracy. Notably, some frontal lobe activations appeared to be primarily in white matter, surpassing a threshold of *p* < .001 uncorrected, which warranted further examination when it appeared again in the between-groups comparison, as described below.

We applied a double subtraction approach to compare BOLD signal activations between the groups. We used the contrast values obtained from the within-groups comparison (first subtraction) to compare differences between groups (second subtraction). Fig 2 shows positive BOLD signals that represented greater “forward minus control” activation differentials in the PTLD group than in the control group (exceptions are noted in Table 2 where activations were higher in the control condition for the control group, yielding a “false” hyperactivation in the PTLD group).

**Fig 2.**
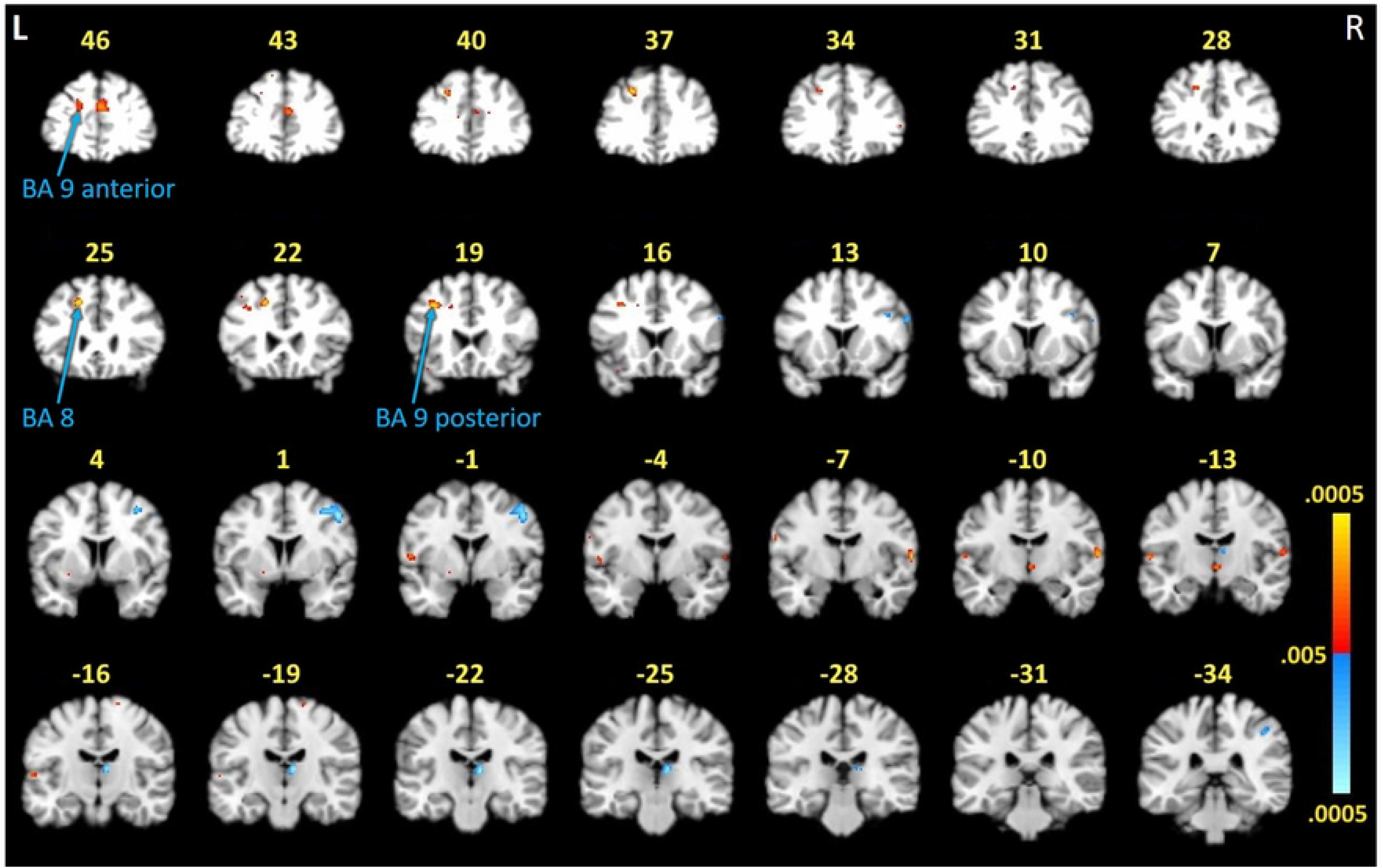
FMRI activation differences between study groups during the working memory task. Activations represent a double subtraction between groups (i.e., the difference between-groups of the difference within-groups [(forward, 2 stimulus) minus (control, 2 stimulus)]). Red indicates activity in PTLD > controls, except for where indicated in Table 2. Blue indicates activity in controls > PTLD. Areas of greater activation in control participants (blue) compared to PTLD participants were consistent with localized activity previously documented as relevant to task performance and reflected hypoactivity in these regions by the PTLD group.[23] Unexpectedly, three frontal lobe activations demonstrated by the PTLD group (red) were located primarily within white matter. Numbers denote *y*-axis on the MNI template. Color scale represents .005 < *p* < .0005. L = left, R = right hemispheres.

**Table 2.** Task-related BOLD activations between PTLD and control participants. Regions are based on Talairach coordinates^27^ and listed anterior-posterior (y-plane). A = Brodmann Area; GM = gray matter; WM = white matter. * = positive BOLD signal indicates greatest activity in the control participants during the control condition, which reversed the BOLD signal interpretation in this double subtraction method. The percentage of WM computed within a cluster is noted in parentheses in the GM/WM column (reporting those with WM > 50%). Cluster information is reported at *p* < .005, uncorrected.

| Cluster size | T-value | x, y, z (MNI) | Brain region | GM/WM | p-value |
| --- | --- | --- | --- | --- | --- |
| **Controls > PTLD** |  |  |  |  |  |
| 17 | 3.14 | 58, 14, 26 | Right inferior frontal gyrus (BA 9) | GM | <.005 |
| 10 | 3.01 | 40, 12, 30 | Right inferior frontal gyrus (BA 9) | GM | <.005 |
| 124 | 3.69 | 48, 0, 44 | Right precentral gyrus (BA 6) | WM (64%) | <.001 |
| 95 | 3.68 | 8, -24, 8 | Right thalamus | GM | <.001 |
| 21 | 3.08 | 44, -36, 44 | Right inferior parietal lobule (BA 40) | GM | <.005 |
| **PTLD > Controls** |  |  |  |  |  |
| 120 | 3.37 | 0, 50, 24 | Left medial frontal gyrus (BA 9) | GM | <.005 |
| 33 | 3.22 | -18, 48, 26 | Left superior frontal gyrus (BA 9) | WM (89%) | <.005 |
| 40 | 3.82 | -22, 38, 36 | Left middle frontal gyrus (BA 8) | WM (41%) | <.001 |
| 56 | 3.97 | -18, 24, 40 | Left middle frontal gyrus (BA 8) | WM (80%) | <.001 |
| 46 | 3.72 | -34, 18, 38 | Left precentral gyrus (BA 9) | WM (99%) | <.001 |
| 26 | 3.31 | -56, -4, 6 | Left precentral gyrus (BA 6) | GM | <.005 |
| 45 | 3.61 | 62, -10, 8 | Right superior temporal gyrus (BA 42)* | GM | <.001 |
| 18 | 3.15 | 2, -14, -4 | Right thalamus | GM | <.005 |
| 23 | 3.33 | -62, -16, 6 | Left superior temporal gyrus (BA 41)* | GM | <.005 |

Increased task-related activity in the controls versus PTLD participants was observed in regions that were consistent with the original study and other similar paradigms, such as the premotor cortex, thalamus, and inferior parietal lobe [23–25, 40]. These results also indicated that group differences were due to the PTLD group showing hypo-activation (or not activating at all) in brain regions normally associated with the task, even though their accuracy was normal.

Increased task-related activity in PTLD participants versus controls was observed in the frontal lobe (BA 8 and 9). While frontal lobe involvement would be expected in this working memory task, the clusters of task-related activation were located primarily within white matter, as opposed to the regions of relative hypoactivation noted above which were predominantly in gray matter. One white matter region was activated more robustly in controls than in the PTLD group (in BA 6). Closer inspection of this region at the more conservative p < .001 threshold, however, indicated that it was actually comprised of two smaller gray matter activations within close proximity that, when smoothed, bridged white matter. Additional analyses were conducted to compute the percentage of white matter included in each ROI for these four frontal lobe activations, as described below. Based on these findings, masks were created for each ROI that allowed us to compute MRI signal contrast values within these circumscribed regions for correlations with DTI and clinical variables (described below).

### Tissue class segmentation results

Calculation of the overlap between the white matter mask derived from the tissue segmentation analysis and the ROIs derived from the fMRI working memory contrast (forward minus control conditions) resulted in the identification of four significant functional activation clusters that were primarily localized to white matter (i.e. over 50% of the voxels within the functional ROI mask overlapped with the white matter tissue segmentation mask). In three frontal activation areas, the patients with PTLD showed elevated activation compared to controls (Fig 3A) localized to the left BA 8 ROI (80.06% white matter), the left BA 9 anterior ROI (89.39% white matter), and the left BA 9 posterior ROI (99.73% white matter). By contrast, in one significant cluster, controls showed increased activation relative to patients localized to the right BA 6 ROI (63.56% white matter). The remaining 10 activation clusters obtained from functional imaging analysis were categorized as non-majority white matter, with eight of the ROIs ranging from 0% - 11.81% white matter and two with 40.63% and 41.30% white matter respectively.

**Fig 3.**
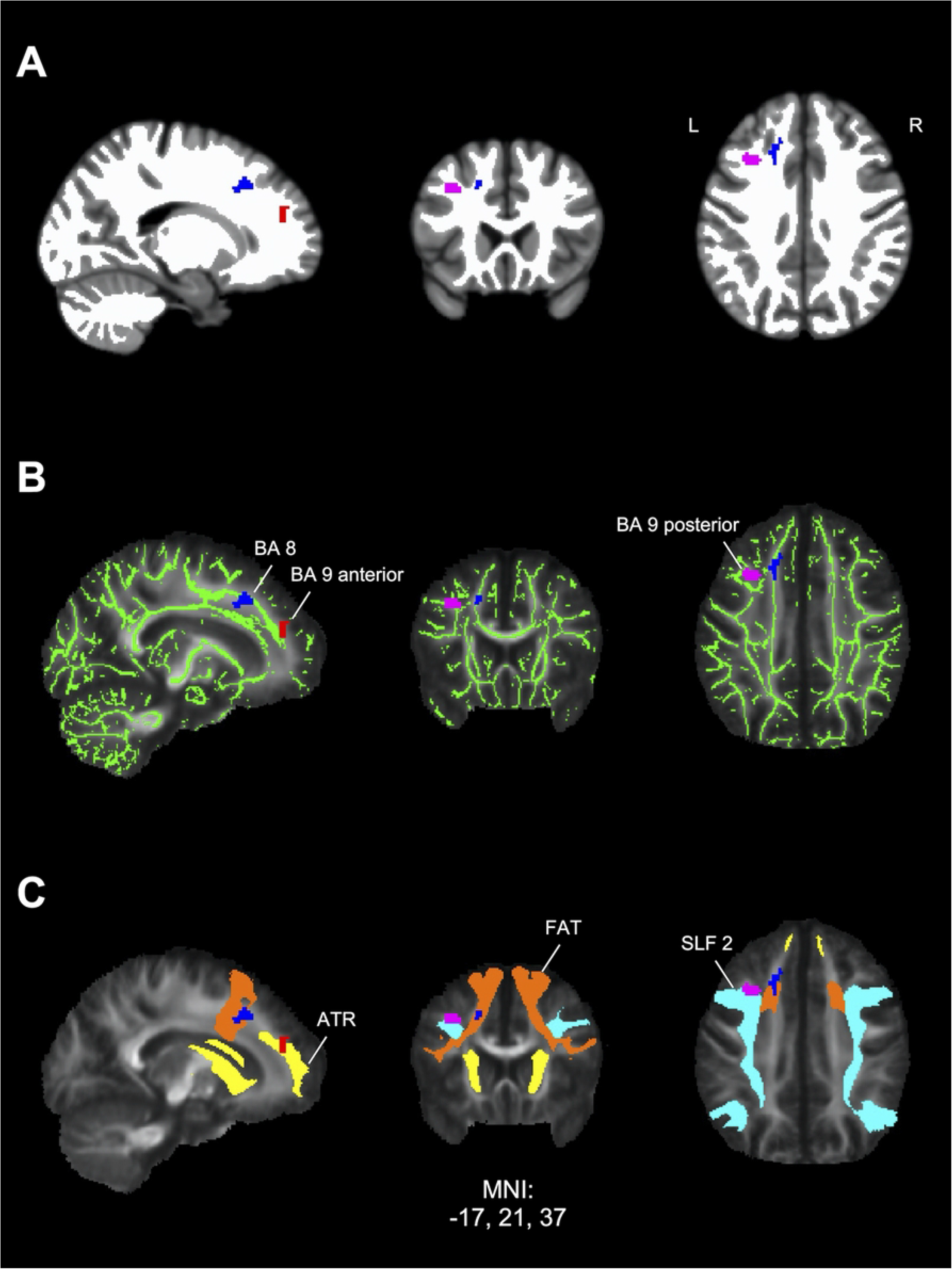
Localization of frontal task-related fMRI activations in white matter. Significant frontal clusters of elevated activation in PTLD participants compared to controls from the contrast values of the forward minus control conditions were transformed into binary regions of interest (ROIs) labeled by their Brodmann area location. Blue: Brodmann area 8 (BA 8) ROI, red: anterior Brodmann area 9 (BA 9 anterior) ROI, pink: posterior Brodmann area 9 (BA 9 posterior) ROI. Labeled ROIs were used to compute the percent overlap with each of the following measures. **(A)** White matter mask (white) derived from a study-specific T1 template using tissue class segmentation analysis. The BA 8 ROI showed 80.06% overlap, the BA 9 anterior ROI showed 89.39% overlap, and the BA 9 posterior ROI showed 99.73% overlap with white matter. **(B)** Mean fractional anisotropy (FA) and white matter skeleton (green) maps derived using diffusion tensor imaging (DTI) analysis showing overlap with fMRI ROIs. The BA 8 ROI showed 25.00% overlap, the BA 9 anterior ROI showed 22.35% overlap, and the BA 9 posterior ROI showed 24.46% overlap with skeletonized white matter. For reference, the white matter skeleton accounts for 34.47% of the overall white matter mask from the tissue class segmentation. **(C)** Mean FA map overlaid with three long range white matter pathways obtained from DTI. The BA 8 ROI showed 41.57% overlap with the frontal aslant tract (FAT; orange), the BA 9 anterior ROI showed 49.62% overlap with anterior thalamic radiation (ATR; yellow), and the BA 9 posterior ROI showed 57.61% overlap with the superior longitudinal fasciculus 2 (SLF 2; cyan). L = left, R = right hemispheres.

### DTI results

A whole-brain between-groups analysis did not show significant group differences (FWE-corrected *p* > .05). However, we were specifically interested in examining the integrity of white matter within the regions identified in the fMRI analysis.

#### Overlap with white matter skeleton

Overlap with the white matter skeleton derived from the TBSS analysis was computed for ROIs obtained from the fMRI analysis that exhibited greater than 50% white matter as determined by the tissue segmentation analysis (see above). The area of task-related activation in the left BA 8 showed 25.00% overlap with the white matter skeleton, while left BA 9 anterior ROI showed 22.35% overlap, and the left BA 9 posterior ROI showed 24.46% overlap (Fig 3B). Overlap with the white matter skeleton was substantially lower for the right BA 6 ROI (13.46%).

#### Overlap with long range white matter tracts

For the three white matter frontal activation ROIs that exhibited elevated task activation in patients, a follow-up analysis was completed to identify long-range fiber pathways that showed overlap with the task-related activation ROIs. The left BA 8 ROI exhibited 41.57% overlap with the frontal aslant tract (FAT, Fig 3C) and 17.70% overlap with the first branch of the superior longitudinal fasciculus (SLF 1). The left BA 9 anterior ROI exhibited 49.62% overlap with the anterior thalamic radiation (ATR, Fig 3C), 12.12% overlap with the dorsal cingulum, and 13.26% overlap with the forceps minor. Finally, the left BA 9 posterior ROI showed 57.61% overlap with the second branch of the superior longitudinal fasciculus (SLF 2, Fig 3C).

#### Relationship between white matter microstructure and duration of illness

Within the PTLD group, an exploratory whole brain analysis was conducted to identify areas that demonstrated a significant relationship between DTI microstructural measures (FA, MD, AD, and RD) and DOI. Significant positive correlations with DOI were observed in right frontal regions (FWE-corrected *p* < .05) for both MD and AD (Fig 4). Across both DTI measures, the correlations were found in regions consistent with the right ATR and SLF 3. In MD and AD, voxels with a significant positive correlation with DOI primarily overlapped with the right ATR (60.55% and 17.73%, respectively) and the right SLF 3 (23.39% and 54.29%, respectively). No significant correlations emerged for FA or RD, and no significant negative correlations were found.

**Fig 4.**
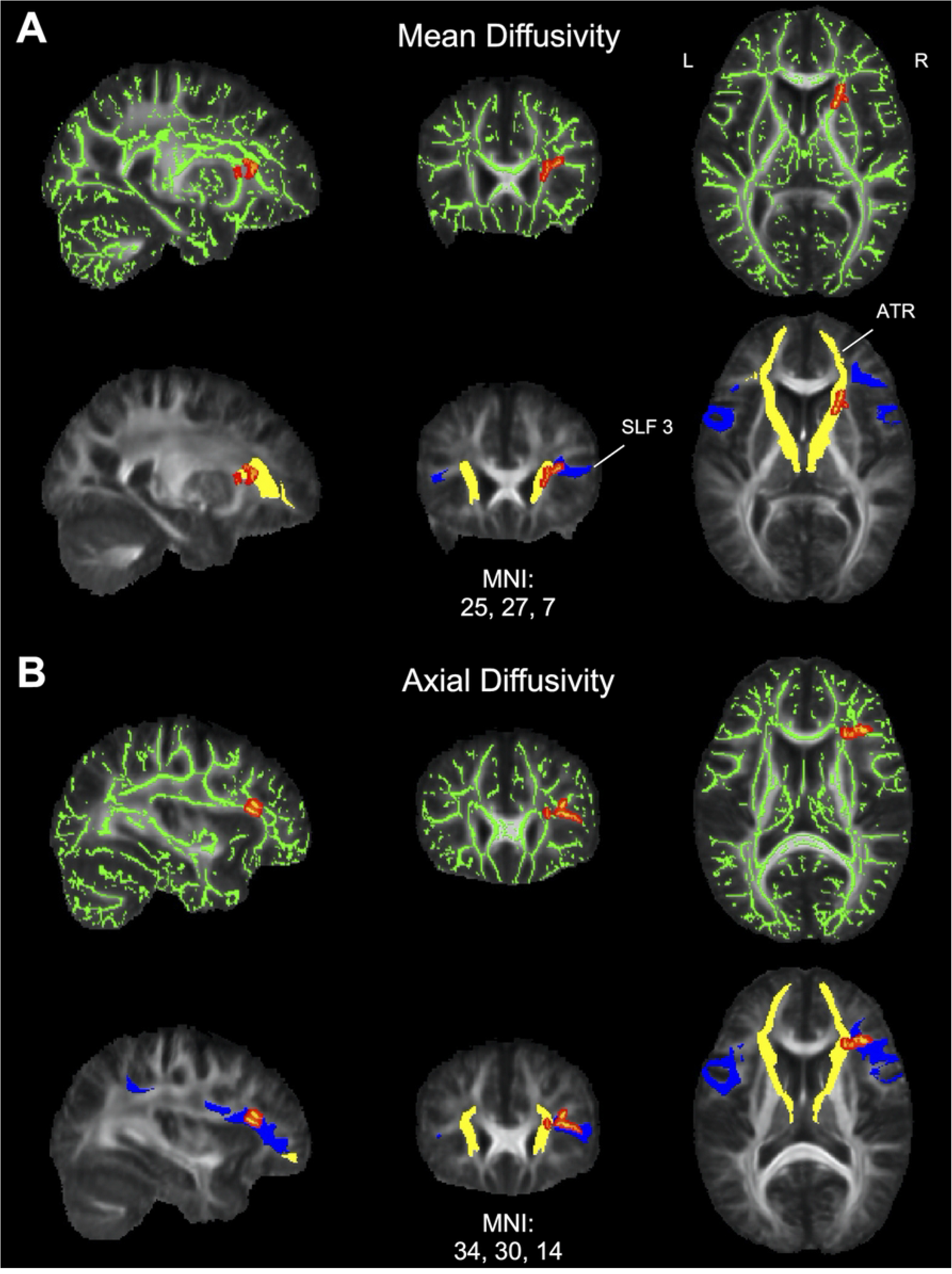
Relationship between diffusion tensor imaging (DTI) metrics and duration of illness (DOI) in PTLD. (A) Mean diffusivity (MD) results. Top: area of significant correlations between MD and DOI overlaid on the mean fractional anisotropy (FA) map and white matter skeleton (green). Positive correlations are displayed in red (FWE-corrected *p* < .05). There were no significant negative correlations.Bottom: MD results overlaid with two long range white matter pathways for visualization showing overlap with anterior thalamic radiation (ATR; yellow) and superior longitudinal fasciculus 3 (SLF 3; blue). (B) Axial diffusivity (AD) results. Top: area of significant correlations between AD and DOI overlaid on the mean FA map and white matter skeleton (green). Positive correlations are displayed in red (FWE-corrected *p* < .05). There were no significant negative correlations. Bottom: AD results overlaid with ATR (yellow) and SLF 3 (blue) showing overlap for localization. L = left, R = right hemispheres.

### Relationship among fMRI, DTI, and clinical variables

In the PTLD group, we explored the relation among the three clusters of activation localized to the white matter and their respective DTI axial diffusivity measures, using fMRI-derived ROI masks applied to the fMRI and DTI skeletonized maps (Fig 5). The fMRI and DTI BA 9 ROIs marginally and negatively correlated, *r*(11) = -.55, *p* = .077 (S3 File Table 3). Thus, greater axial diffusivity was tentatively associated with less white matter activation in this region.

**Fig 5.**
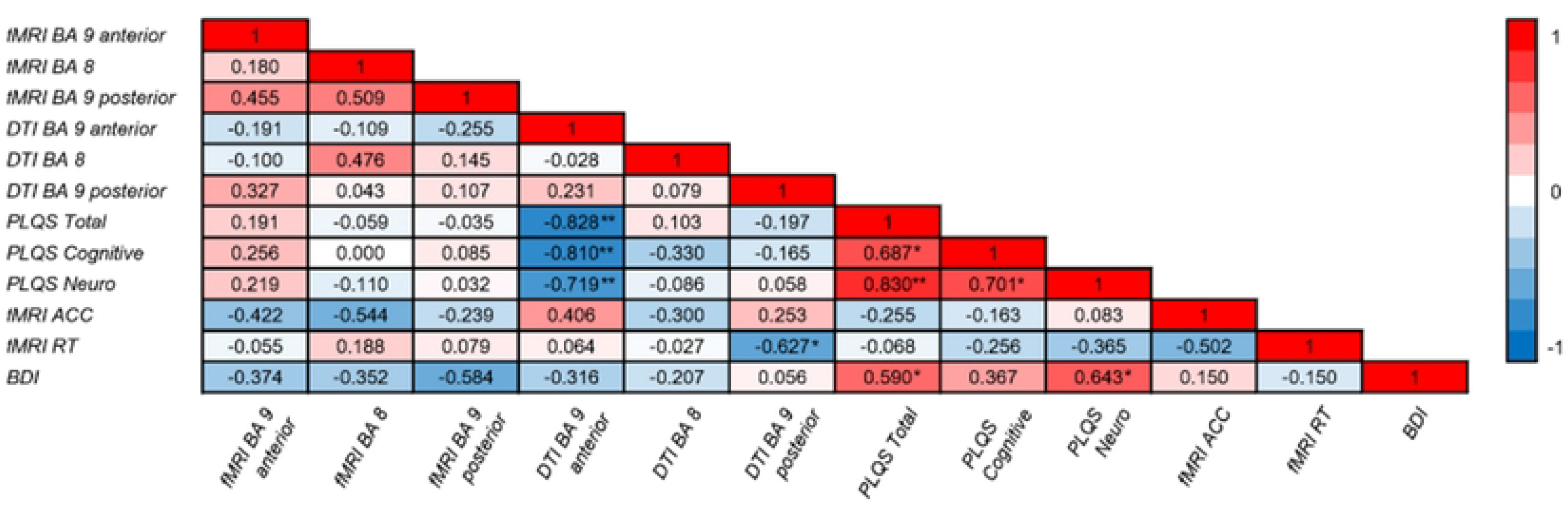
Correlation matrix of the relationship among fMRI, DTI, and clinical variables. Brain regions correspond to the regions of interest revealed by fMRI between-groups contrasts that were located in white matter. FMRI beta values and DTI axial diffusivity measures were correlated with clinical variables. Notably, DTI measures of the BA 9 anterior region negatively correlated with PLQS scores (i.e., higher axial diffusivity was associated with fewer symptoms). PLQS = post-Lyme questionnaire of symptoms, fMRI ACC = accuracy on the fMRI task, fMRI RT = response times on the fMRI task, BDI = Beck Depression Inventory; * = *p* ≤ .05; ** = *p* ≤ .01 (two-tailed).

DTI skeletonized axial diffusivity within the three ROIs were correlated with the sum of symptoms on the PLQS (total, cognitive, and neurologic) to assess the clinical relevance of these regions. Axial diffusivity in the BA 9 anterior ROI negatively correlated with all three clinical measures (total: *r*(12) = - .76, *p* = .004; neurological: *r*(12) = -.78, *p* = .003; cognitive: *r*(12) = -.81, *p* = .001), showing that greater diffusivity was associated with *<u>fewer</u>* symptoms (Fig 5). The two other ROIs did not correlate with sum of symptoms [all *p*-values > .29] (S3 File Table 4). A similar comparison between the fMRI beta weight contrast values within these ROIs and the sum of PLQS measures also did not correlate [all *p*-values > .45] (S3 File Table 4). Thus, these associations point to frontal lobe axial diffusivity, specifically, as a potential indicator of healthy outcomes in PTLD (i.e., higher axial diffusivity with fewer symptoms).

We also compared the three ROIs from fMRI beta contrast and DTI axial diffusivity values to accuracy and RT performance on the working memory task. FMRI values and DTI values did not correlate with accuracy or response times [all *p*-values > .10] (S3 File Table 4). Axial diffusivity in these three regions also did not correlate with accuracy [all *p*-values > .22] (S3 File Table 4). Higher axial diffusivity in the left BA 9 posterior ROI correlated with lower (faster) response times, Spearman’s *r*(11) = -.63, *p* = .039 [the two other *p*-values > .85].

Given that depressive symptoms can accompany PTLD [7, 41], we probed for the influence of depressive symptoms on fMRI, DTI, and clinical variables using the Beck Depression Inventory (BDI) total score [42]. The BDI total score correlated only with fMRI BA 6 beta values, Spearman’s r(11) = - .767, p = .006, and did not correlate with any other fMRI ROI values (gray or white matter) or axial diffusivity measures [all *p*-values > .05] (S3 File Table 4). However, a higher BDI total score positively correlated with the total sum of symptoms, Spearman’s *r*(12) =.59, *p* = .044 and sum of neurological symptoms, Spearman’s *r*(12), *p* = .024 [but not with cognitive symptoms, *p* = .24]. The BDI total score did not correlate with accuracy or response times on the working memory task [both *p*-values > .66] (S3 File Table 4).

Taken together, these associations suggest that increased axial diffusivity impacts clinical variables in a positive way. Notably, white matter axial diffusivity may be a marker of healing during PTLD and represent a healthier outcome.

## Discussion

This study applied multimodal neuroimaging methods to examine brain structure and function in a carefully selected sample of people with well-characterized PTLD in the absence of co-morbid diseases and other factors that could otherwise explain the results. The original hypothesis that the PTLD group would show altered task-related activations, as revealed by fMRI, was supported. The PTLD group activated different gray brain regions relative to controls, some of which were not previously associated with this task in a prior study of healthy adults.[23] Moreover, the PTLD group hypoactivated other areas that were relevant to the task, relative to controls, suggesting that the PTLD group relied on compensatory mechanisms to complete the task, given that they performed as well as controls did.

Unexpectedly, three of the PTLD group’s activated regions found in the frontal lobe were located in white matter. Event-related white matter findings are distinctly unusual in fMRI studies because of the relatively low energy demands and blood volume in white matter [43–46]. These factors render the white matter BOLD signal comparably much less detectable than in gray matter. However, evidence has accumulated in recent years suggesting that hemodynamic changes can be detected in white matter using high magnetic field strength fMRI methods [45, 47]. Explanations for the BOLD signal detection in white matter, especially in an event-related manner, range from reduced physiological noise relative to signal, to visualization of action potentials, to the energy associated with neurovascular coupling of astrocytes [48]. It is notable that the oxygen extraction fraction in white matter has been reported to be comparable to that of gray matter, a finding that may be explained by the need to maintain resting membrane potentials in white matter oligodendrocytes [46, 49]. Thus, white matter changes may signify dysfunction- or excessive function-of glial cells [2]. Because this study was not originally designed to examine white matter function, additional studies are needed to further examine the white matter activations observed here.

We scrutinized further the locations of three white matter frontal lobe fMRI-guided ROIs using tissue segmentation analyses. Results showed that the left BA 9 anterior, BA 8, and BA 9 posterior ROIs contained 80% or more overlap with white matter tissue. Overlaying these three ROIs onto a skeletonized DTI map, which is a highly conservative mapping that accounts for 34% of the overall white matter mask, indicated 22% or more intersection. These results confirmed that task-related activity can be localized to the white matter of the frontal lobe in people with PTLD.

These three ROIs were subsequently examined in more detail exploring their white matter structural integrity using DTI methods (i.e., diffusivity measures). We found that, as a group, the PTLD participants’ DTI measures did not differ from that of controls. When we compared clinical and neurologic symptoms measured outside of the scanner environment to DTI measures in the PTLD group, we found that greater axial diffusivity was associated with fewer symptoms. Thus, the white matter DTI changes observed in this study may represent a healthy marker of the neurological repair process.

We found that axial and mean diffusivity increased with DOI in regions within the right frontal lobe. It should be noted that DOI *per se* did not correlate with clinical measures from the PLQS, behavioral measures from the fMRI task, or BDI scores (all p-values > .42), indicating that diffusivity measures were the driving factor behind the correlations. The regions identified by the diffusivity/DOI correlation were discrete from the white matter ROIs derived from the fMRI task, which were instead located in the left frontal lobe. Importantly, both measures implicated white matter changes within the frontal lobe. Cognitive difficulties referable to the frontal lobe are commonly reported in patients with PTLD [7, 13, 50].

The measure of axial diffusivity is thought to be related to axonal properties, such as diameter, count, and density [51–55]. However, it is important to note that the literature reflects ambiguity with respect to whether increased or decreased axial diffusivity is related to axonal injury. Some studies have reported axonal damage associated with axial diffusivity increases [56–59], while others have reported axonal damage associated with axial diffusivity decreases [60]. Moreover, studies have shown that axial diffusivity patterns can differ by region of interest [61–64]. While it is not possible to fully elucidate the biological basis of the altered diffusion signal in the data reported here, our findings demonstrate an important link between axial diffusivity and clinical outcomes, as well as DOI.

The relationship of these unexpected white matter findings to the clinical features of PTLD suggest that white matter abnormalities may have an important role in the symptomatology of PTLD. Prior studies have reported white matter abnormalities in PTLD. Notably, Fallon *et al*. (2003) reported brain perfusion abnormalities in white matter regions, including increased blood flow to the frontal lobe [18]. The current findings supported those of Fallon *et al*. using DTI methods that measured white matter integrity, guided by fMRI activations. The convergence of data across imaging modalities underscore the role and vulnerability of the frontal lobe in PTLD symptoms.

This study was limited by the small sample size. First, this sample size potentially introduced a risk of false positive findings by limited statistical power. However, several measures were employed to mitigate this risk by independently confirming our results in multiple ways. We identified white matter activations that passed a threshold of p < .001, uncorrected (Table 2 and S2 Table), a commonly used threshold for fMRI data reporting. We confirmed that the white matter activations from the fMRI analyses were, indeed, located in white matter using a follow-up segmentation analysis. We then created localized regions of interest specifically from these fMRI-guided activations and used these regions exclusively in our correlations with clinical variables. Given that these regions passed several thresholds, using independent multimodal imaging approaches, the converging findings suggest plausible results in this relatively small sample size.

Second, generalization of these findings to the larger PTLD community is also mitigated by the relatively small sample size and stringent inclusion criteria (e.g., exclusion of atypical early presentation of Lyme disease that did not meet CDC criteria). The homogeneity of the demographics within our sample further limited generalizability of the findings, given that the study included people with a relatively advanced education level, high socioeconomic status, residing within the mid-Atlantic region of the United States. Third, due to the small sample size, we could not statistically control for the time between infection and antibiotic treatment, antibiotic dose, and duration of treatment across participants. Such factors may have influenced healing and recovery processes that could not be fully accounted for in this study. Fourth, this was a cross-sectional study with a wide ranging DOI across participants. Ideally, participants would be followed longitudinally from disease onset in order to better monitor the changes associated with PTLD directly over time. Nonetheless, given the careful selection of participants with PTLD and little else to medically explain these results, these data provide an important preliminary look at structural and functional brain changes associated with PTLD and guide future neurologically-based research in the field.

This study represents an in-depth examination of the integrity of brain structure and function in people with PTLD using more sophisticated neuroimaging measures than has been reported to date. The findings provide quantitative, objective measures of brain changes that can be associated with clinical and cognitive measures. Importantly, these findings support and validate PTLD patient reports of cognitive difficulties [7]. Results reported here may have implications for other diseases in which white matter pathology has been demonstrated (e.g., multiple sclerosis) or in illnesses in which cognitive complaints follow disease onset in the absence of objective methods to confirm neuropathology (e.g., chronic fatigue syndrome, fibromyalgia, post acute COVID) [65–68]. The use of multimodal neuroimaging methods, like the ones used in the current study, may be a viable approach for obtaining information on brain function and structure to identify biomarkers of disease burden.

## Data Availability

All data and analysis code are available on the Open Science Framework (https://osf.io/kshq7).

## Acknowledgements

We thank Bronte Wen for assistance with fMRI data processing and Cheryl Novak and Susan Joseph for the recruitment of study participants.

## Supporting information

**S1 Table. FMRI task BOLD activations within healthy controls.** Brain regions are based on Talairach coordinates. Bold indicates activations that overlapped with a prior study in young, healthy adults using the same fMRI task by Marvel & Desmond, 2012. All regions met threshold criteria for *p* < .001, uncorrected.

**S2 Table. FMRI task BOLD activations within PTLD.** Brain regions are based on Talairach coordinates. Bold indicates activations that overlapped with a prior study in young, healthy adults using the same fMRI task by Marvel & Desmond, 2012. ^ = regions that were primarily located in white matter. All regions met threshold criteria for *p* < .001, uncorrected.

**S3 File Table 1. FMRI, DTI, and Clinical Variable Descriptive Statistics.** Data are shown as: mean (standard deviation), 95% confidence interval [lower limit, upper limit]. Shapiro-Wilk tests were used for tests of normality. Significant values of the Shapiro-Wilk tests are denoted in bold, p ≤ .05, two-tailed.

**S3 File Table 2. Gray Matter fMRI and Beck Depression Inventory (BDI) Descriptive Statistics.** Data are shown as: mean (standard deviation), 95% confidence interval [lower limit, upper limit]. Shapiro-Wilk tests were used for tests of normality. Significant values of the Shapiro-Wilk tests are denoted in bold, p ≤ .05, two-tailed.

**S3 File Table 3. Correlations Between fMRI ROIs and Respective DTI ROIs.** Pearson’s tests were used for bivariate correlations involving BA 9 values because they have a normal distribution. A Spearman’s test was used for bivariate correlations involving BA 8 values because they had a non-normal distribution.

**S3 File Table 4. Correlations Between fMRI and DTI ROIs and fMRI Task Accuracy, RT, Symptoms, and BDI.** Spearman’s tests were used for bivariate correlations involving Task Accuracy, RT, Cognitive Symptoms, and BDI because they had a non-normal distribution. Pearson’s tests were used for bivariate correlations involving Total Clinical Symptoms and Neurological Symptoms because they had a normal distribution.

## Notes

### Competing Interest Statement

The authors have declared no competing interest.

### Funding Statement

Funding for this project was generously provided by an anonymous donor to JNA. The funders had no role in study design, data collection and analysis, decision to publish, or preparation of the manuscript.

### Author Declarations

The IRB of Johns Hopkins University School of Medicine gave ethical approval for this work.

### Summary of Updates

upload of the 3 supplement files

