## Supplemental Table 1 for "A multimodal neuroimaging study of brain abnormalities and clinical correlates in post treatment Lyme disease"

**S1 Table**

**FMRI task BOLD activations within healthy controls**

| **Cluster size** | ***T*-value** | ***x*, *y*, *z* (MNI)** | **Brain region (BA)** |
| --- | --- | --- | --- |
| **(*n*=18)** 20 \| 4.26 \| -44, 44, 14 \| L Middle frontal gyrus (BA 10); 193 \| 7.16 \| **40, 36, 30** \| R Superior frontal gyrus (BA 9); 201 \| 4.71 \| **-32, 18, 2** \| L Anterior insula (BA 13); 680 \| 8.05 \| **36, 14, 2** \| R Anterior insula (BA 13); 2451 \| 6.44 \| **12, 14, 44** \| R Middle frontal gyrus (BA 32); 1044 \| 5.38 \| **-48, 4, 22** \| L Precentral gyrus (BA 6); 18 \| 3.83 \| **10, 2, 20** \| R Caudate; 31 \| 5.09 \| -8, -12, 76 \| L Superior frontal gyrus (BA 6); 2598 \| 6.11 \| **6, -24, 12** \| Thalamus; 4558 \| 7.53 \| **44, -38, 42** \| R Inferior parietal lobule (BA 44); 77 \| 5.56 \| -50, -50, -12 \| L Inferior temporal gyrus (BA 20); 347 \| 4.79 \| **-32, -62, -28** \| L Cerebellar crus I; 1315 \| 5.68 \| **36, -68, -22** \| R Cerebellar crus I; 114 \| 4.96 \| 28, -68, 6 \| R Posterior cingulate (BA 30); 59 \| 4.18 \| -22, -76, -50 \| L Cerebellar crus II **Brain regions are based on Talairach coordinates.** Bold indicates activations that overlapped with a prior study in young, healthy adults using the same fMRI task by Marvel & Desmond, 2012. All regions met threshold criteria for *p* < .001, uncorrected. \| \| | | | |
