## Supplemental Table 2 for "A multimodal neuroimaging study of brain abnormalities and clinical correlates in post treatment Lyme disease"

| **S2 Table**  **FMRI task BOLD activations within PTLD**  **Cluster size *T*-value *x, y, z* (MNI) Brain region (BA)**  **(*n*=11)** | | | |
| --- | --- | --- | --- |
| 36 | 5.17 | **-34, 32, 28** | L Middle frontal gyrus (BA 9) |
| 270 | 6.47 | **-26, 30, 4** | L Inferior frontal gyrus (BA 45) |
| 87 | 5.66 | 28, 30, 30 | R Middle frontal gyrus (BA 9)^ |
| 11 | 4.52 | -20, 26, 28 | L Medial frontal gyrus (BA 9)^ |
| 12 | 4.49 | **30, 24, 6** | R Anterior insula (BA 13) |
| 214 | 5.45 | **2, 22, 48** | Supplementary motor area |
| 18 | 5.86 | **18, 18, 14** | R Caudate |
| 73 | 7.24 | 24, 16, 34 | R Middle frontal gyrus (BA 8)^ |
| 14 | 5.04 | -26, 14, 38 | L Middle frontal gyrus (BA 8)^ |
| 20 | 4.68 | **28, 8, 52** | R Middle frontal gyrus (BA 6) |
| 21 | 6.92 | -18, 4, -10 | L Putamen |
| 113 | 5.12 | -14, 4, 44 | L Cingulate gyrus (BA 24)^ |
| 7 | 4.36 | **-32, -2, 58** | L Middle frontal gyrus (BA 6) |
| 133 | 6.15 | -26, -6, 42 | L Middle frontal gyrus (BA 6) |
| 52 | 7.22 | 26, -24, -4 | R hippocampus |
| 18 | 5.48 | 10, -44, 0 | R Parahippocampus |
| 10 | 4.92 | -8, -46, -4 | L Cerebellar lobe IV |
| 9 | 4.62 | -22, -52, 30 | White matter, undefined region^ |
| 573 | 6.6 | 20, -56, 22 | R Precuneus (BA 31) |
| 133 | 12.49 | -22, -58, 4 | L Lingual gyrus (BA 18) |
| 9 | 5.04 | -30, -70, -54 | L Cerebellar crus II |
| 20 | 4.86 | **6, -76, 40** | R Precuneus (BA 7) |
| 39 | 5.22 | **-24, -78, 40** | L Precuneus (BA 19) |
| **Brain regions are based on Talairach coordinates.** Bold indicates activations that overlapped with a prior study in young, healthy adults using the same fMRI task. ^ = regions that were primarily located in white matter. All regions met threshold criteria for *p* < .001, uncorrected. | | | |
