## Supplemental File 3 for "A multimodal neuroimaging study of brain abnormalities and clinical correlates in post treatment Lyme disease"

**Table 1: FMRI, DTI, and Clinical Variable Descriptive Statistics**

|  | PTLD | Controls |
| --- | --- | --- |
| fMRI ROI:  BA 8  BA 9 anterior  BA 9 posterior | .009 (.012), 95% [.002, .016], W = .94, p = .52  .009 (.017), 95% [-.002, .021], W = .96, p = .73  .017 (.020), 95% [.004, .031], W = .95, p = .66 | -.007 (.012), 95% [-.013, -.001], W = .94, p = .34  -.010 (.016), 95% [-.018, -.002], W = .95, p = .45  -.006 (.016), 95% [-.013, .002], W = .93, p = .20 |
| Axial diffusivity ROI:  BA 8  BA 9 anterior  BA 9 posterior | .001 (.00008), 95% [.001, .001], W = .97, p = .93  .001 (.00006), 95% [.001, .001], W = .94, p = .43  .001 (.00002), 95% [.001, .001], W = .95, p = .63 | .001 (.00008), 95% [.001, .001], W = .83, **p = .019**  .001 (.00008), 95% [.001, .001], W = .98, p = .99  .001 (.0001), 95% [.001, .001], W = .89, p = .11 |
| PLQS:  Total  Neurologic  Cognitive | 8.17 (4.39), 95% [5.38, 10.95], W = .94, p = .48  4.50 (2.023) 95% [3.21, 5.79], W = .90, p = .16  2.00 (1.35), 95% [1.14, 2.86], W = .71, **p = .001** | ---------- |
| FMRI task  *Accuracy:*  Control condition  1 stimulus  2 stimuli  Forward condition  1 stimulus  2 stimuli  *Response time:*  Control condition  1 stimulus  2 stimuli  Forward condition  1 stimulus  2 stimuli | 97.8 (2.00), 95% [96.4, 99.1], W = .79, **p = .008**  97.2 (2.60), 95% [95.4, 98.9], W = .72, **p = .001**  94.9 (10.0), 95% [88.2, 101.6], W = .55, **p < .001**  89.8, (12.0), 95% [88.2, 101], W = .70, **p = .001**  1102 (326), 95% [883, 1321], W = .87, p = .075  1226 (417), 95% [946, 1506], W = .75, **p = .002**  1221 (455), 95% [946, 1527], W = .84, **p = .034**  1800 (632), 95% 1375, 2225), W = .83, **p = .025** | 96.9 (5.02), 95% [94.4, 99.4], W = .66, **p < .001**  95.7 (6.35), 95% [92.5, 98.8], W = .71, **p < .001**  96.0 (3.69), 95% [94.2, 97.8], W = .84, **p = .007**  91.3 (4.75), 95% [89.0, 93.7], W = .94, p = .26  922 (184), 95% [831, 1014], W = .98, p = .94  1011 (198), 95% [913, 1110], W = .89, **p = .039**  996 (217), 95% [888, 1104], W = .98, p= .97  1373 (384), 95% [1182, 1564], W = .96, p = .64 |

Data are shown as: mean (standard deviation), 95% confidence interval [lower limit, upper limit]. Shapiro-Wilk tests were used for tests of normality. Significant values of the Shapiro-Wilk tests are denoted in bold, p < .05, two-tailed.

**Table 2: Gray Matter fMRI and Beck Depression Inventory (BDI) Descriptive Statistics**

|  | PTLD |
| --- | --- |
| fMRI ROI:  Left Medial Frontal Gyrus (BA9)  Left Precentral Gyrus (BA6)  Right Thalamus | .019 (.041), 95% [-.008, .047], W = .97, p = .916  .009 (.021), 95% [-.005, .023], W = .93, p = .452  .014 (.028), 95% [-.005, .032], W = .88, p = .090 |
| BDI Total Score | 18.42 (12.05), 95% [10.76, 26.07], W = .79, **p = .008** |

Data are shown as: mean (standard deviation), 95% confidence interval [lower limit, upper limit]. Shapiro-Wilk tests were used for tests of normality. Significant values of the Shapiro-Wilk tests are denoted in bold, p < .05, two-tailed.

**Table 3: Correlations Between fMRI ROIs and Respective DTI ROIs**

| *ROIs* | *DTI BA 8* | *DTI BA 9 anterior* | *DTI BA 9 posterior* |
| --- | --- | --- | --- |
| fMRI BA 8 | r(11) = .44, p = .18 |  |  |
| fMRI BA 9 anterior |  | **r(11) = -.55, p = .077** |  |
| fMRI BA 9 posterior |  |  | r(11) = .11, p = .76 |

Pearson’s tests were used for bivariate correlations involving BA 9 values because they have a normal distribution. A Spearman’s test was used for bivariate correlations involving BA 8 values because they had a non-normal distribution.

**Table 4: Correlations Between fMRI and DTI ROIs and fMRI Task Accuracy, RT, Symptoms, and BDI**

| *Region of Interest (ROI)* | *Task Accuracy* | *Task Reaction Time (RT)* | *Total Clinical Symptoms* | *Neurological Symptoms* | *Cognitive Symptoms* | *Beck Depression Index Total Score (BDI)* |
| --- | --- | --- | --- | --- | --- | --- |
| fMRI BA 8 | r(10) = -.54  p = .10 | r(10) = .18  p = .60 | r(11) = .19  p = .57 | r(11) = .22  p = .52 | r(11) = .26  p = .45 | r(11) = -0.352  p = 0.289 |
| fMRI BA 9 Anterior | r(10) = -.42  p = .22 | r(10) = -.055  p = .88 | r(11) = -.059  p = .86 | r(11) = -.11  p = .75 | r(11) = .000  p = 1.00 | r(11) = -0.374  p = 0.257 |
| fMRI BA 9 Posterior | r(10) = -.24  p = .51 | r(10) =.07  p = .83 | r(11) = -.035  p = .92 | r(11) =.032  p = .93 | r(11) = .085  p = .80 | r(11) = -0.584  p = 0.059 |
| fMRI Left Medial Frontal Gyrus (BA9) | r(10) = -.30  p = .40 | r(10) = .07  p = .86 | r(11) = -.12  p = .72 | r(11) = -.097  p = .78 | r(11) = -.085  p = .80 | r(11) = -0.584  p = 0.059 |
| fMRI Left Precentral Gyrus (BA6) | r(10) = -.20  p = .58 | r(10) = .02  p = .96 | r(11) = -.006  p = .99 | r(11) = .012  p = .97 | r(11) = -.021  p = .95 | **r(11) = -0.767**  **p = 0.006** |
| fMRI Right Thalamus | r(10) = -.31  p = .39 | r(10) =.15  p = .68 | r(11) = .097  p = .78 | r(11) = -.01  p = .978 | r(11) = .23  p = .49 | r(11) = -0.151  p = 0.658 |
| DTI BA 8 | r(11) = -.30  p = .37 | r(11) = -.027  p = .94 | r(12) = .10  p = .75 | r(12) = -.086  p = .79 | r(12) = -.33  p = .29 | r(11) = -0.207  p = 0.519 |
| DTI BA 9 Anterior | r(11) = .41  p = .22 | r(11) = .064  p = .85 | **r(12) = -.76**  **p = .004** | **r(12) = -.78**  **p = .003** | **r(12) = -.81**  **p = .001** | r(11) = -0.316  p = 0.317 |
| DTI BA 9 Posterior | r(11) = .25  p = .45 | **r(11) = -.63**  **p = .039** | r(12) = -.20  p = .54 | r(12) =.058  p = .86 | r(12) = -.17  p = .61 | r(11) = 0.056  p = 0.862 |
| Beck Depression Index Total Score (BDI) | r(11) = .15  p = .66 | r(11) = -.15  p = .66 | **r(12) = .59**  **p = .044** | **r(12) = .64**  **p = .024** | r(12) = .37  p = .24 | ------- |

Spearman’s tests were used for bivariate correlations involving Task Accuracy, RT, Cognitive Symptoms, and BDI because they had a non-normal distribution. Pearson’s tests were used for bivariate correlations involving Total Clinical Symptoms and Neurological Symptoms because they had a normal distribution.
